# Measles incidence in South Africa: a six-year review, 2015 - 2020

**DOI:** 10.1101/2022.02.03.22270382

**Authors:** Mukhlid Yousif, Heather Hong, Susan Malfeld, Sheilagh Smit, Lillian Makhathini, Tshepo Motsamai, Dipolelo Tselana, Morubula Manamela, Mercy Kamupira, Elizabeth Maseti, Kennedy Otwombe, Kerrigan McCarthy, Melinda Suchard

**Author notes:** **Corresponding author** Dr Mukhlid Yousif, Centre for Vaccines and Immunology, National Institute for Communicable Diseases, Researcher, Virology department, Faculty of Health Sciences, University of the Witwatersrand.

## Abstract

In 2012 the World Health Organization (WHO) aimed to eliminate measles in five regions by 2020. This retrospective descriptive study reviewed measles surveillance data in South Africa for the period 2015 - 2020 to document the epidemiology of measles and the progress made towards meeting the 2020 measles elimination goal.

A total of 22,578 specimens were tested over the period 2015 - 2020 yielding 401 (1.8%) confirmed measles cases, 321 (1.4%) compatible and 21,856 (96.8%) discarded cases. The most affected age group was 0-4 year olds. At the provincial level, South Africa achieved adequate surveillance, defined as more than two cases of febrile rash notified annually per 100 000 popoulation, except for KwaZulu-Natal and Limpopo in 2020, probably due to COVID-19 lockdown restrictions. Of confirmed cases, only 26% were vaccinated, 3% were too young to receive vaccines, 5% were not vaccinated, and 65% had vaccination status unknown. Measles vaccine effectiveness amongst 1-4 year olds was 80%. Using the standard case definition, South Africa achieved the measles elimination target of less than one case per one million nationally in years 2015, 2016 and 2020. The years 2017 to 2019 had incidence rates exceeded one per million nationally. Using a narrow case definition, that excluded positive rubella cases, improved the indicators with only the year 2017 having an incidence rate of more than one per million.

South Africa displays intermittent measles outbreaks approximately six-yearly interspersed by inter-epidemic periods in which the country meets measles elimination targets. Intense effort is needed to increase the vaccine coverage to avoid periodic outbreaks. Enhanced molecular testing of each case will be required as measles incidence declines regionally.

## Introduction

Measles is a highly contagious airborne disease that affects the upper respiratory tract. Measles is caused by the rubeola virus, a member of the *Morbillivirus* genus, *Paramyxoviridae* family (1). Transmission occurs through direct contact with infectious droplets or by airborne spread when an infected person breathes, coughs, or sneezes. After exposure, the first sign of measles is usually a high fever, followed by runny nose, cough and rash (2). In young children less than five years, around 30% of measles infections can lead to complications such as diarrhoea, otitis media, pneumonia, encephalitis, seizures and death (3).

Before the development of a measles vaccine in the 1960s, measles was a leading cause of morbidity and mortality (4). Measles was responsible for more than two million deaths annually (5). Despite the availability of the vaccine, measles remains a leading cause of death in children under five years of age (6). Measles outbreaks still occur in countries where vaccination coverage is low (7). According to the WHO, in 2018, more than 140,000 people died due to measles, of which at least one third were in Africa (7). In 2012, the WHO updated the measles elimination initiative aiming to eliminate measles by 2020 in at least five of six global regions (8). The WHO defined the elimination of measles as the absence of indigenous measles cases in a certain geographic region for up to 12 months in the presence of a high-quality surveillance system (ref?). The WHO also requires national measles vaccination coverage of 95% in all districts with two doses of measles vaccine per child. At least 80% of districts should investigate one or more suspected cases within a year and should report a non-measles rash illness rate of at least two cases per 100, 000 nationally (9).

In South Africa, the measles vaccine is available in single antigen formulation in the public sector or in combination format with mumps and rubella antigens (MMR) in private sector. In 1975, the measles vaccine was first administered as one dose at nine months of age. In 1995, when the immunization programme was expanded, a second dose was added at 18 months. In 2016, the schedule changed to earlier administration at 6 and 12 months of age (10). Post the introduction of the expanded programme of immunization, several measles outbreaks have occurred. Between 2003 and 2005, an outbreak occurred with 1,676 cases reported (11). In 2009-2010, a large outbreak occurred with 18,431 documented cases (12). In 2017, a small outbreak occurred with measles cases detected in Western Cape, Gauteng and Kwazulu-Natal provinces, with a total number of 186 infected (10). In 2019, a cluster of measles infection in four siblings who travelled to Georgia was detected in Cape Town (13). Correspondingly, between 2012 and 2017 the measles 1^st^ dose measles vaccination coverage in South Africa averaged 71.7%, while measles 2^nd^ dose vaccination averaged 68.8% (14). Since then, measles 2^nd^ dose coverage increased to 76.4% in 2018 but remains below the 95% coverage level required for elimination, thus sporadic cases still occur.

As part of febrile rash surveillance, any suspected case of measles seen by a clinician should be notified within 24 hours and a blood specimen should be collected and sent to the National Institute for Communicable Diseases (NICD) for testing. Febrile rash cases thus comprise multiple aetiologies, the common of which in the South African setting is rubella. In this manuscript we review the febrile surveillance data for the period 2015 to 2020, to document the epidemiology of measles in South Africa, and the progress made towards national measles elimination. Rubella incidence has been previously reported (Hong et al, submitted)

## Methods

### Study design

A retrospective descriptive study was conducted to review measles surveillance data in South Africa for the period 2015-2020 to document the epidemiology of measles and the progress made towards meeting the 2020 measles elimination goal. Ethical approval was obtained from the Human Research Ethics committee of the Unversity of the Witwatersrand, under: : “essential communicable disease surveillance and outbreak investigation activities of the National Institute for Communicable Disease of (NICD) of South Africa”.

### Case-based Surveillance

For all suspected measles cases meeting the case definition of febrile rash with at least one of the symptoms; cough, coryza or conjunctivitis, or in any patient in whom a clinician suspected measles, a case investigation form (CIF) was required to be filled and sent to the NICD along with a serum sample. Throat swab and/ or urine samples were not routinely collected during this period.

### Specimen testing

All serum samples were tested for measles immunoglobulin M (IgM) and rubella IgM using a commercial Enzyme-Linked Immunosorbent Assay (ELISA) according to manufacturer instructions. For the period 2015 to 2017, sera were tested using Enzygnost® kits (Siemens AG, Erlangen, Germany). For the period 2018 to 2020, sera were tested using Euroimmun® kits (Euroimmun AG, Luebeck, Germany). A second specimen was requested following any equivocal results for measles IgM. From the period 2017 to 2020, all sera that tested positive or equivocal for measles IgM were also tested for the presence of measles virus genome by RT-PCR. Ideally, the specimens of choice for measles RT-PCR are throat swabs and urine specimens, however, sera occasionally yield positive results. Measles genotyping to differentiate wild from vaccine strains was conducted for any RT-PCR positive samples (RT-PCR cycle threshold (CT) value <35). Genotyping was performed by amplifying 450 nucleotides of the nucleocapsid region followed by sequencing and phylogenetic analysis (15).

### Case Classification

Based on the laboratory and epidemiological investigations, suspected measles cases were classified as follows: (i) discarded, when the case did not meet the clinical or laboratory definition (measles IgM negative, vaccine-associated [within five weeks of measles vaccine, or had vaccine strain present]) (ii) compatible, when the case met the clinical case definition, was not epidemiologically linked, but no blood specimen was received, or blood specimen was equivocal (iii) confirmed, when the case was laboratory-confirmed (measles IgM positive and/or PCR positive). In this study we only report on the laboratory-confirmed cases and do not further discuss the compatible cases, due to the heterogeneous nature of febrile rash aetiology in years with no large measles outbreaks.

In South Africa rubella rubella virus is endemic and rubella vaccination is not part of the expanded programme of immunization. The most common cause of febrile morbilliform rash in our setting is therefore rubella. Cross-reactive measles serology is well described, where measles IgM may be falsely elevated during intercurrent infection with rubella (16). Due to overlapping clinical symptoms, such cases are usually classified as both “confirmed measles” and “confirmed rubella” cases due to the difficulty of excluding a measles diagnosis and the need to err on the side of caution regarding early measles outbreak response. We have therefore reported our laboratory confirmed measles cases using two definitions – firstly all laboratory-confirmed measles cases (standard definition as per WHO guidelines), secondly after exclusion of cases that were dual positive for rubella IgM (narrow definition).

### Data analysis

Data were captured and analyzed in Microsoft Excel 2016. A descriptive analysis was performed. Categorical data were reported as frequencies or percentages, while continuous data were reported as median and interquartile range (IQR).

### Vaccine effectiveness

Vaccine effectiveness was determined using the narrow case definition to exclude confounding by rubella cases. Vaccine effectiveness was calculated among 1-4 year olds, only due to predominantly missing vaccine information in older age groups. Vaccine efficacy (VE) was estimated using the formula VE=((ARU-ARV)/ARU) * 100 where ARU was the measles attack rate in the unvaccinated population and ARV was the measles attack rate in the vaccinated population. Factors associated with measles infection were determined by univariate and multivariate logistic regression. Analysis was conducted for cases occurring up to 2016 and after 2016, due to the vaccine schedule change that occurred in 2016. Statistical analysis was conducted using SAS Enterprise Guide 7.15 (SAS Institute, Cary, NC, USA) assuming a 0.05 level of significance.

## Results

Of the 22,578 patients tested over the period 2015 - 2020, 11,179 (49.5%) were males, 10,782 (47.8%) were females and 617 (2.7%) had unknown sex. The median age was 5.0 (IQR 3.0-8.0) years. Measles IgM tested positive in 465 (2.1%) samples, equivocal in 433 (1.9%) and negative for 21,386 (94.7%) samples. Over the period between 2017 to 2020, 454 real-time PCR tests were performed, of which 143 (31.5%) were positive. Among the PCR positive cases, 40 specimens were subjected to a genotyping assay, of which 39 were determined as genotype D8 and one specimen was genotype B3, which was an imported case from Saudi Arabia (10).

Of the total of 899 cases that tested positive or equivocal for measles IgM and/or positive for measles PCR, 401 (44.6%) were classified as laboratory-confirmed measles cases, 321 (35.7%) were compatible, and 166 (18.5%) were discarded. Of the confirmed cases 28.9% (116/401) also tested positive for rubella IgM.

Figure 1. shows the trend of confirmed measles cases over the six years (2015 - 2020). Measles cases ranged from 0 to 14 per month, with exception of 2017, in which confirmed cases ranged from 3 to 28 per month, corresponding with an outbreak (**Figure 1A**). After excluding rubella positive cases, measles cases ranged from 0 to 7 per month except in 2017 (**Figure 1B**). Measles cases occurred mostly in the age group of 0-4 years (n=148, 37.0%), and 20-44 years (n=105, 26.2%) (**Figure 2**). The age-standardized incidence rates showed that the group 0-4 years had the highest incidence rates (**Table 1**). The sex distribution among the age groups did not show any significant pattern.

**Table 1:** Age-specific incidence rates of laboratory-confirmed measles in South Africa, 2015-2020.

| Age group | Measles cases | Population | Rate per million population |  |
| --- | --- | --- | --- | --- |
|  |  |  | Standard case definition<br>(Including dual positive for rubella | Narrow case definition<br>(Excluding positive rubella |
| 2015 |  |  |  |  |
| 0_4 | 14 | 5,936,350 | 2.4 | 2.2 |
| 5_9 | 1 | 5,537,225 | 0.2 | 0.0 |
| 10_14 | 1 | 5,138,468 | 0.2 | 0.2 |
| 15_19 | 0 | 5,124,373 | 0 | 0.0 |
| 20_44 | 3 | 21,822,066 | 0.1 | 0.1 |
| >45 | 2 | 11,398,439 | 0.2 | 0.2 |
| Total | 21 | 54,956,921 | 0.4 | 0.3 |
| 2016 |  |  |  |  |
| 0_4 | 5 | 5,862,896 | 0.9 | 0.9 |
| 5_9 | 3 | 5,761,111 | 0.5 | 0.3 |
| 10_14 | 0 | 5,183,234 | 0 | 0.0 |
| 15_19 | 1 | 4,873,874 | 0.2 | 0.2 |
| 20_44 | 8 | 22,659,494 | 0.4 | 0.3 |
| >45 | 1 | 11,568,256 | 0.1 | 0.1 |
| Total | 18 | 55,908,865 | 0.3 | 0.3 |
| 2017 |  |  |  |  |
| 0_4 | 51 | 5,866,573 | 8.7 | 7.8 |
| 5_9 | 31 | 5,76,4576 | 5.4 | 3.5 |
| 10_14 | 18 | 5,093,681 | 3.5 | 3.3 |
| 15_19 | 22 | 4,592,001 | 4.8 | 4.4 |
| 20_44 | 73 | 23,439,277 | 3.1 | 3.1 |
| >45 | 8 | 11,765,840 | 0.7 | 0.7 |
| **Total** | **203** | **5,652,1948** | **3.6** | **3.2** |
| **2018** |  |  |  |  |
| 0_4 | 41 | 5,928,951 | 6.9 | 3.0 |
| 5_9 | 14 | 5,862,081 | 2.4 | 0.3 |
| 10_14 | 0 | 5,252,485 | 0 | 0.0 |
| 15_19 | 0 | 4,733,790 | 0 | 0.0 |
| 20_44 | 9 | 23,681,676 | 0.4 | 0.3 |
| >45 | 1 | 12,266,622 | 0.1 | 0.1 |
| **Total** | **65** | **57,725,605** | **1.1** | **0.5** |
|  |  |  | Standard case definition<br>(Including dual positive for rubella) | Narrow case definition<br>(Excluding positive rubella) |
| 2019 |  |  |  |  |
| 0_4 | 31 | 5,733,946 | 5.4 | 1.9 |
| 5_9 | 12 | 5,737,439 | 2.1 | 0.0 |
| 10_14 | 5 | 5,427,902 | 0.9 | 0.4 |
| 15_19 | 4 | 4,660,002 | 0.9 | 0.9 |
| 20_44 | 12 | 24,137,303 | 0.5 | 0.2 |
| >45 | 3 | 13,078,429 | 0.2 | 0.2 |
| Total | 67 | 5,877,5021 | 1.1 | 0.4 |
| 2020 |  |  |  |  |
| 0_4 | 6 | 5,743,450 | 1.0 | 0.7 |
| 5_9 | 13 | 5,715,952 | 2.3 | 0.5 |
| 10_14 | 2 | 5,591,553 | 0.4 | 0.4 |
| 15_19 | 1 | 4,774,579 | 0.2 | 0.2 |
| 20_44 | 0 | 24,418,106 | 0.0 | 0.0 |
| >45 | 0 | 13,378,710 | 0.0 | 0.0 |
| Total | 22 | 59,622,350 | 0.4 | 0.2 |

Blocks shaded in grey show incidence rates that exceeded one case per million population. Population figures as per mid-year population estimates 2020 (statistics South Africa, 2020). Measles cases shown in column two were all laboratory-confirmed.

**Figure 1:**
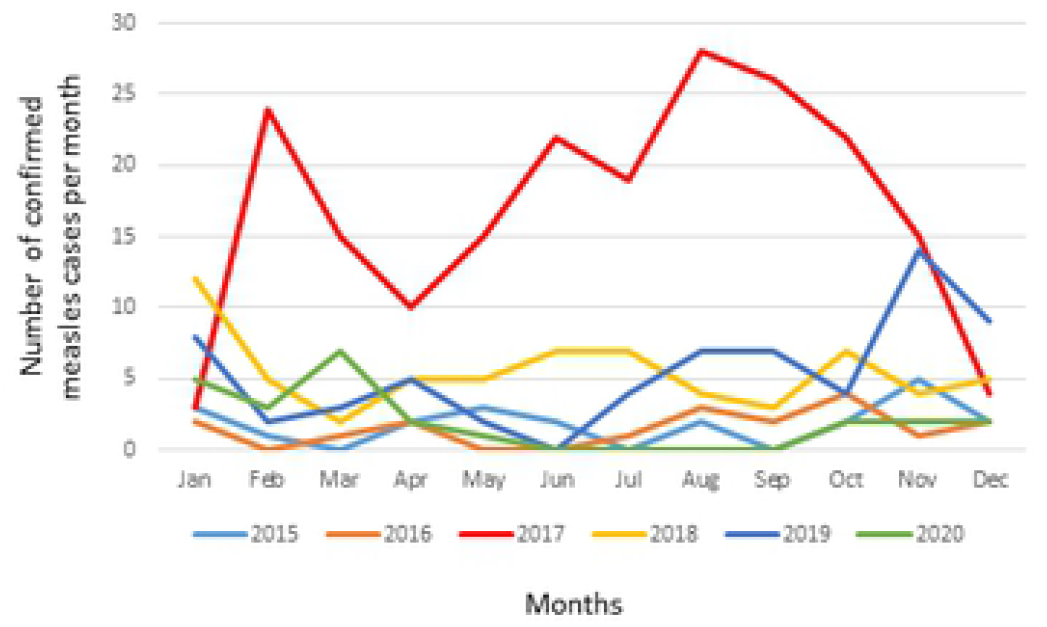
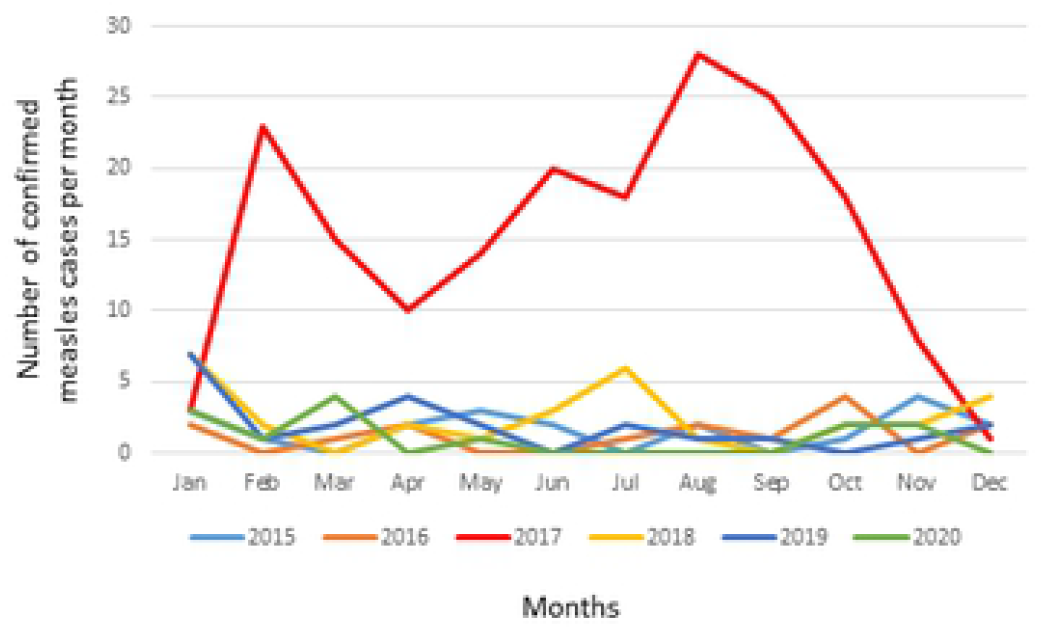
Monthly incidence of measles infection in South Africa during 2015-2020. **Figure 1A** shows confirmed measles cases including rubella positive cases. **Figure 1B** shows confirmed measles cases excluding rubella cases.

**Figure 2:**
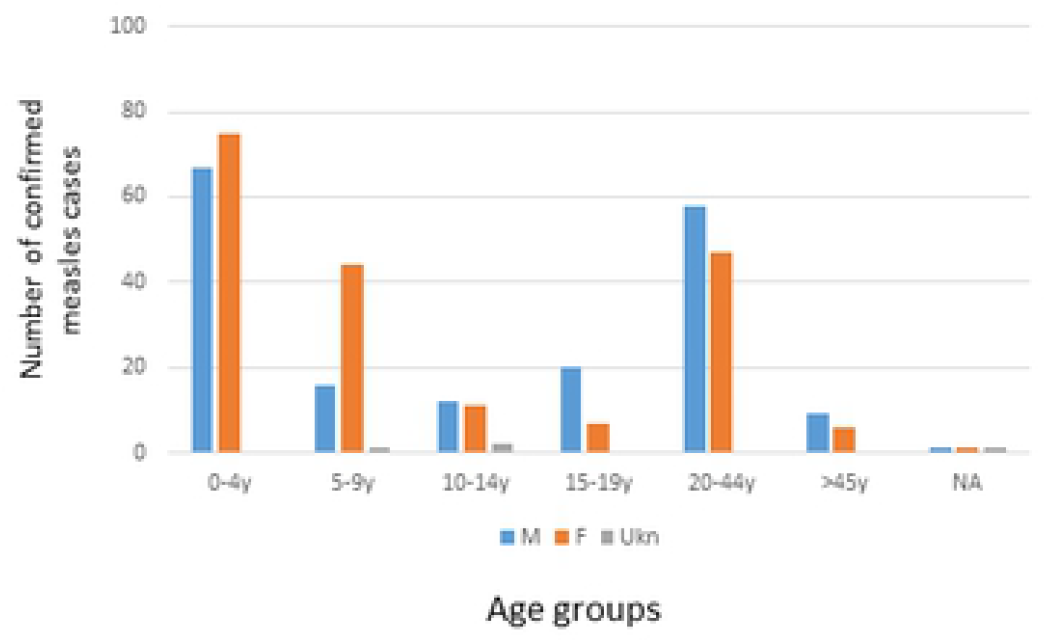
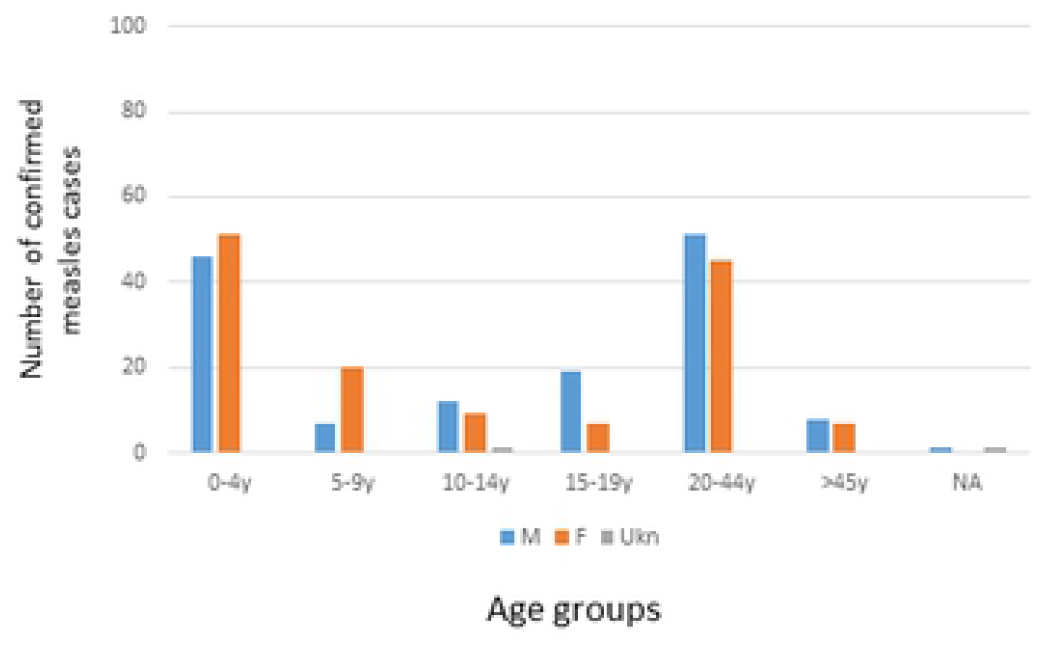
Age and sex distribution among measles cases identified during 2015-2020. **Figure 2A** includes cases dual positive for rubella IgM. **Figure 2B** shows cases excluding rubella IgM positive cases.

Gauteng province had the highest number of confirmed measles cases (n=141, 35.2%) followed by KwaZulu-Natal (n=95, 23.7%), Western Cape (n=67, 16.7%), Eastern Cape (n=28, 7.0%), North West (n=26, 6.5%), Free State (n=13, 3.2%), Mpumalanga (n=11, 2.7%), while Northern Cape, and Limpopo had the least number of cases (n=10, 2.1%). To understand the provincial incidence rates we calculated the number of cases per million population by using the population midyear estimates (17) (**Table 2**).

**Table 2:**
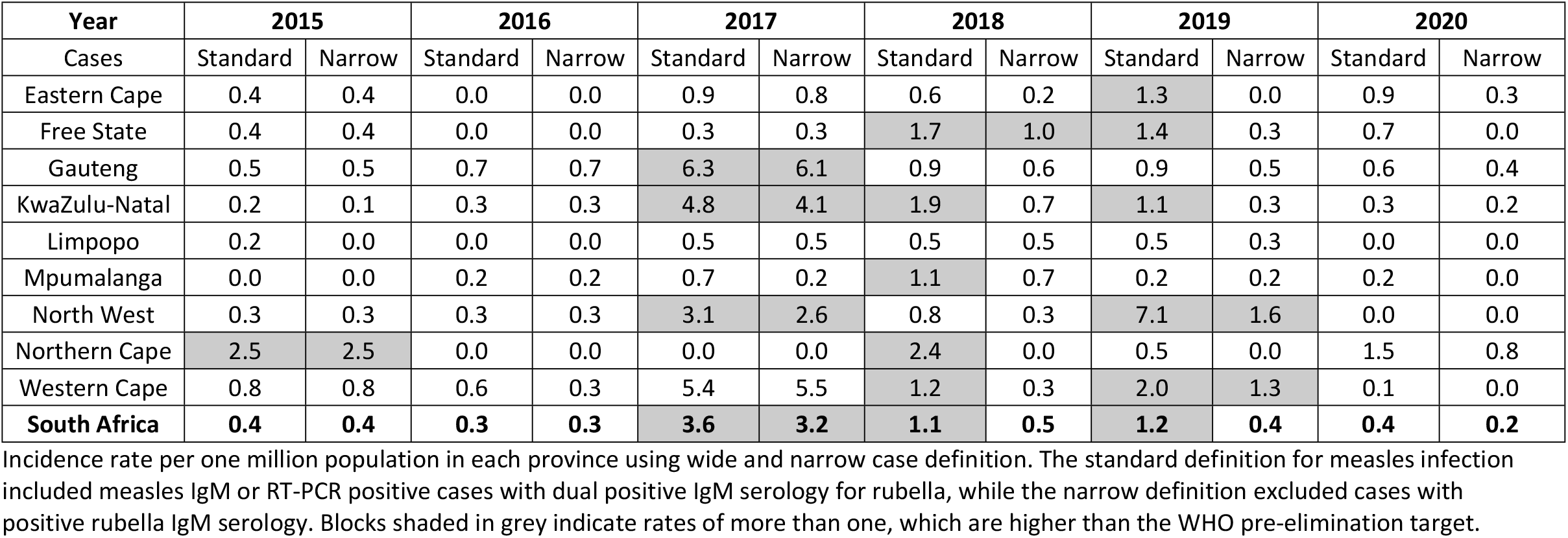
Provincial incidence rates of laboratory-confirmed measles during 2015-2020.

The WHO elimination goal of less than one measles case per one million population was achieved in each province in 2015, 2016 and 2020, except in Northern Cape in 2015. However, in the years 2017, 2018 and 2019 many provinces had more than one case per million (**Table 2**). Repeating the same analysis excluding cases in which rubella IgM was dual positive yielded incidence rates above one per million in 2017 in most provinces, and in Free State in 2018, North West and Western Cape in 2019 (**Table 2**).

According to the WHO, surveillance adequacy should be measured by the number of non-measles febrile rash illness cases reported per 100,000 population. More than two cases per 100,000 population is required for adequate surveillance. Using this indicator, South Africa achieved adequate surveillance indicator target throughout 2015 – 2020, except in KwaZulu-Natal and Limpopo in 2020, corresponding to the lockdown imposed due to the COVID-19 restrictions (**Table 3**).

**Table 3:**
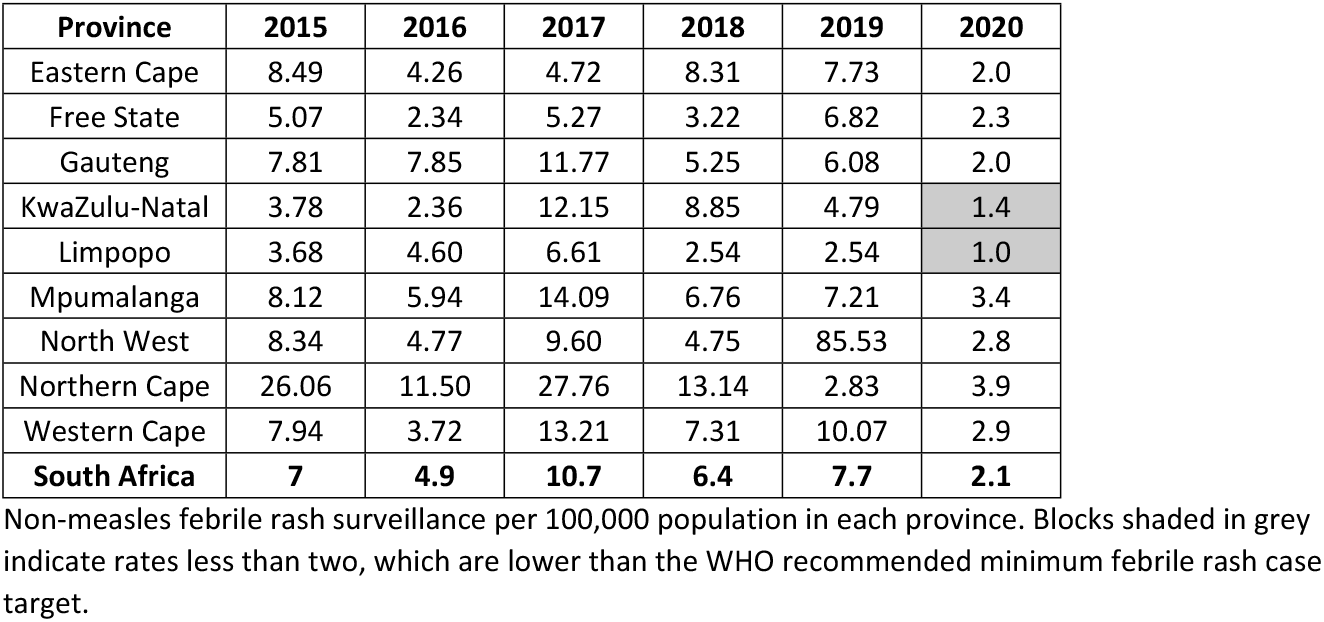
Surveillance adequacy per province during 2015-2020.

Additional indicators showed only 105 (26%) of laboratory-confirmed cases were measles vaccinated, 22 (5%) were not measles vaccinated, 14 (3%) were too young (< 6 months) for measles vaccination, and measles vaccination status of 260 (65%) were unknown. Among the group in which measles vaccination was reported, 25 (24%) received only one dose, 53 (50%) had two or more doses, and 28 (27%) had an unknown number of doses. Repeating this analysis after exclusion of cases who were dual positive for rubella IgM, only 45 (16%) were vaccinated, of which 17 (41%) had two doses. Among measles negative samples, measles vaccination status was unknown in 13887 (63%) of cases. CIFs were submitted with specimens in 192 (48%) cases, unique EPID numbers were submitted in 204 (51%) cases, and only 141 (35%) cases had both CIF and unique EPID number (**Table 4**).

**Table 4:** Surveillance indicators for laboratory-confirmed measles and non-measles cases.

| Status |  | Confirmed measles cases<br>(standard definition)<br>(n=401) | % | Confirmed measles cases<br>(narrow case definition)<br>(n=285) | % | non-measles<br>(22,177) | % |
| --- | --- | --- | --- | --- | --- | --- | --- |
| Measles vaccination | Too young <6 m | 14 | 3 | 7 | 2 | 713 | 3 |
|  | Unknown | 260 | 65 | 219 | 77 | 13,887 | 63 |
|  | Yes | 105 | 26 | 45 | 16 | 7,327 | 33 |
|  | No | 22 | 5 | 14 | 5 | 250 | 1 |
| Measles vaccine doses | 1 | 25 | 24 | 19 | 42 | 726 | 10 |
|  | 2 or more | 53 | 50 | 17 | 38 | 4,628 | 63 |
|  | Unknown | 28 | 27 | 5 | 11 | 1,972 | 27 |
| Case investigation form |  | 192 | 48 | 118 | 41 | 10,853 | 49 |
| Epidemiological number |  | 204 | 51 | 124 | 44 | 11,343 | 51 |
| Case investigation form and epidemiological number |  | 141 | 35 | 84 | 29 | 7,444 | 34 |

Standard case definition included all laboratory-confirmed (IgM positive or PCR positive) measles cases, including those dual positive for rubella IgM. Narrow case definition included all laboratory-confirmed (IgM positive or PCR positive) measles cases, excluding those dual positive for rubella IgM. Non-measles cases are febrile rash cases that tested negative for measles.

Over the period of 2015 - 2020, South Africa failed to meet the WHO recommendation for immunization coverage target of children under one-year-old. South Africa had less than 95% coverage in all provinces over the period 2015-2019 except Gauteng in 2015. Vaccination coverage was below 90% in three districts in South Africa between November 2020 and January 2021 (19). Of note, vaccination coverage was the lowest in 2017, corresponding with the 2017 measles outbreak (18).

Of the 22,587 febrile rash cases, there were 8,127 (36.0%) aged 1-4 years old with the majority being females (**Table 5**).

**Table 5:**
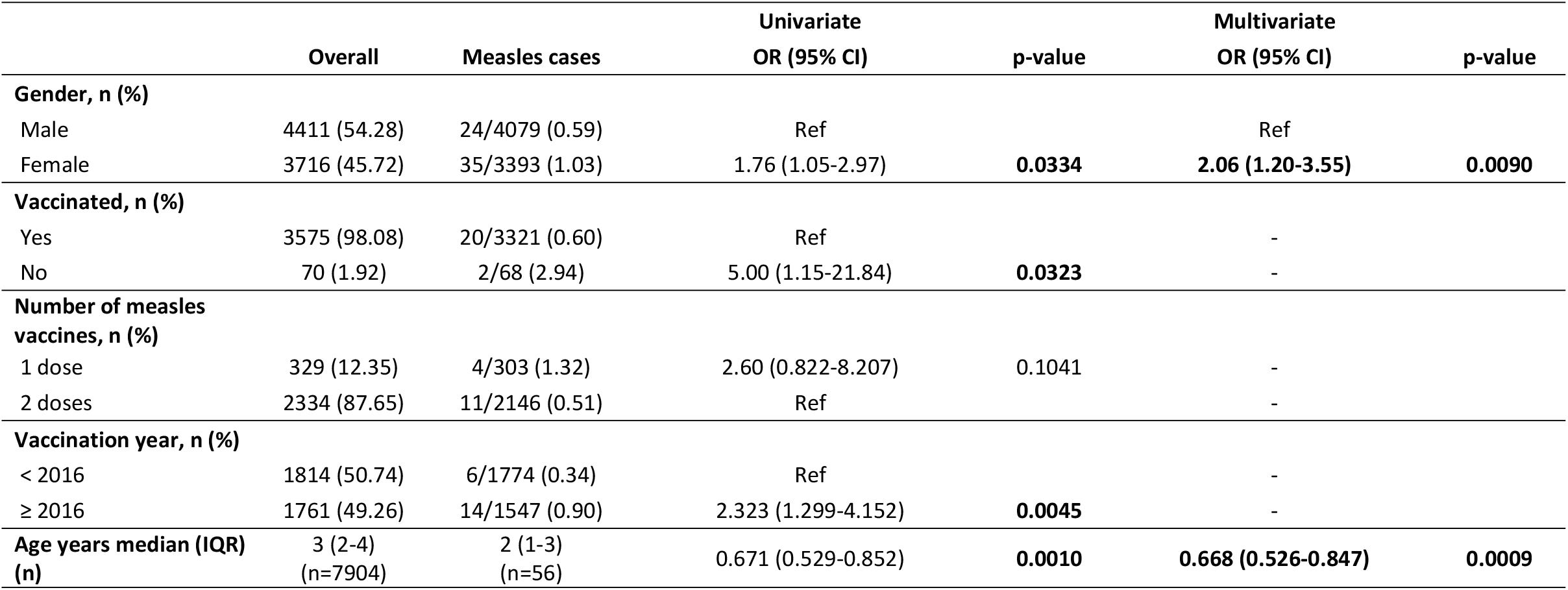
Factors associated with measles diagnosis among children 1-4 years old.

Overall, the median (IQR) age of febrile rash cases was 3 (2–4) years whereas that of those with measles was 2 (1–3) years. Multivariate logistic regression showed that compared to males, females had a higher odds of measles infection (OR: 2.06, 95% CI: 1.20-3.55, p=0.009) whereas each year of age reduced the odds of infection (OR: 0.67, 95% CI: 0.53-0.85, p=0.0009). The measles vaccine effectiveness among 1-4 year olds was 80%. On univariate analysis, the odds of measles cases being unvaccinated compared with vaccinated was 5.00 (95%CI: 1.15-21.84, p=0.0323) although measles vaccination status was no longer a significant predictor of measles infection on multivariate analysis. Using univariate analysis, the probability of infection with measles after vaccination was higher after 2016 compared to the previous programme before 2016 (p=0.0045), although the vaccination year was no longer significant on multivariate analysis.

## Discussion

In this review, we aimed to evaluate the level of South Africa’s readiness to eliminate measles. We reviewed six years’ retrospective data from the national surveillance programme for febrile rash illness. Between the years 2015 to 2020, a total of 285 confirmed measles cases (excluding rubella infections) were detected in South Africa, with the highest incidence rate of 6.1 cases per million detected in 2017 in Gauteng province, while the lowest incidence rate of infection was zero detected in many provinces in multiple years (**Table 2**).

Younger children aged from 0-4 years were the most affected age group. Stratified by population figures, the highest incidence rate was in the age group of 0-4 years at 7.8 per million in 2017, 3.0 per million in 2018, and 1.9 per million in 2019, however in 2017 all age groups had had high incidence rates, with many adult cases, due to the outbreak that affected the country (**Table 1**). In 2017, one death was reported (ref?) but outcome data for most cases was not available.

In the past six years, South Africa had a good surveillance system in place, evidenced by the adequate non-measles rash surveillance rate of more than 2.0 per 100,000 population in all provinces from 2015 to 2020, except in 2020 in KwaZulu-Natal and Limpopo, which had a rate of 1.4 and 1.0 per 100,000 respectively. This reduction of the rate of non-measles rash surveillance was probably due to the lockdown from March to August imposed because of the COVID-19 pandemic, resulting in reduced health-seeking behavior but also lowering the transmission of respiratory-borne viruses. The non-measles rash surveillance in South Africa in 2020 was 2.1 per 100,000, still above the recommended threshold.

On the other hand, certain indicators were poorly performed such as the completion of CIF, assignment of unique EPID number, and completion of vaccination information of confirmed and discarded measles cases. In addition, the national immunization coverage of vaccination in all provinces did not reach 95% coverage (18). Imperfect vaccine coverage explains the circulation of measles cases.

Interestingly, choice of the measles case definition plays an important role in evaluating the status of South Africa’s measles elimination goals. Using the standard case definition, South Africa only achieved the elimination target of an incidence rate of less than one case per one million nationally in the years 2015, 2016 and 2020. The years 2017 to 2019 had incidence rates greater than one per million nationally. Conversely, using a narrow case definition that excluded positive rubella cases from the analysis improved the indicators. Only the year 2017 had an incidence rate of more than one per million. In years 2018 and 2019 South Africa kept the incidence rate below one, which means the country is approaching achieving the measles elimination goals. In the year 2020, all provinces had had a rate below one per million populations, which could be explained by COVID-19 restrictions and interruption of the spread of respiratory illnesses generally due to social distancing, increased hygiene measures and lockdowns, or by hesitation in seeking medical services during lockdown periods.

While we cannot definitively conclude that all dual positive measles and rubella samples were due to rubella rather than measles, rubella is more common in the South African setting, and therefore the positive predictive value of a positive rubella IgM result is higher than the positive predictive value of a positive measles IgM result. With relatively few measles cases diagnosed, additional confirmatory tests are required to confirm measles positive results, thus our results excluding the dual positive cases is likely the more accurate estimate.

Conducting measles and rubella serology on all samples is a strength of our study and allowed us to differentiate between samples positive only for measles and those positive for both measles and rubella. Cross-reactive serology occurs reasonably uncommonly using the ELISA methodology, however, the influence of false-positive serology can form a large proportion of cases when overall measles numbers are low and rubella numbers high. Of note, cross-reactive measles and rubella serology has been reported with most commercial assays (16) (20)(21) and likely represents biological increases in polyclonal antibody titres in patients *in vivo*, rather than *in vitro* flaws of the diagnostic kits. Such challenges indicate the need for improved molecular diagnostics for routine measles surveillance in South Africa, necessitating future collection of throat swabs, urine samples or other suitable samples for confirmatory measles molecular testing. Rubella vaccine introduction to South Africa is likely within the next few years and may alleviate testing ambiguities.

Using the narrow case definition, which excluded the rubella positive cases, measles vaccine effectiveness in South Africa was determined as 80% among children aged between 1-4 years old. This is low compared to other studies that reported vaccine effectiveness of 95%, using large datasets (22) (23). Our results also showed that the odds of being vaccinated and having measles was hifher prior to 2016, when children received vaccine at 9 months and 18 months, compared to post 2016 when children receive the vaccine at 6 months and 12 months. Early vaccination might blunt the immune response to subsequent measles vaccine doses (24). Ongoing evaluation of vaccine effectiveness with the new schedule is warranted. A confounder of these results may be that the measles cases in our dataset occurred mostly during the 2017 outbreaks, resulting in most measles cases in our dataset occurring after 2017. Our work is also limited by missing information in our programmatic data, particularly the number of respondents with available information on measles vaccination status and of doses. Nevertheless, our program data provides a good reflection of the impact of the vaccine program in a routine setting.

In conclusion, the results of this study suggested that more effort is needed to increase the vaccine coverage in the country as well as the completion of indicators including clinical investigation form, unique EPID number and information on vaccination status in febrile rash cases. Improvements in laboratory confirmatory measles diagnostic assays will also be required to meet the goals for measles elimination. Moreover, catch-up vaccinations will be needed to fill the gaps particularly following the COVID-19 pandemic in which many children missed their routine immunizations.

## Data Availability

All relevant data are within the manuscript and its Supporting Information files.

## Authors contributions

MY: performed data curation, performed analysis,wrote manuscript, edit and reviewed manuscript. HH: co-conceplized study, performed data curation, reviewed manuscript. SM: performed data curation, reviewed manuscript. SS: performed data curation, reviewed manuscript. LM: performed data curation, review manuscripted. TM: performed data curation, reviewed manuscript. DT: performed data curation, reviewed manuscript. MM: performed data curation, reviewed manuscript. MK: performed data curation, reviewed manuscript. EM: performed data curation, reviewed manuscript. KO: performed analysis, reviewed manuscript. KM: performed analysis, reviewed manuscript. MS: co-conceplized study, performed analysis, reviewed manuscript.

## Acknowledgements

We acknowledge the support and contribution by the provincial expanded programme on Immunisation coordinators, and the provincial communicable disease control teams, notifiable medical conditions teams. We also thank the staff of Center for Vaccines and Immunology at the NICD.

## References

1. Rima BK, Duprex WP. Morbilliviruses and human disease. J Pathol A J Pathol Soc Gt Britain Irel. 2006;208(2):199–214.

2. Battegay R, Itin C, Itin P. Dermatological Signs and Symptoms of Measles: A Prospective Case Series and Comparison with the Literature. Dermatology [Internet]. 2012;224(1):1–4. Available from: https://www.karger.com/DOI/10.1159/000335091

3. CDC. Measles [Internet]. 2020. Available from: https://www.cdc.gov/vaccines/pubs/pinkbook/meas.html#measles

4. Moss WJ. Measles. Lancet [Internet]. 2017;390(10111):2490–502. Available from: http://www.sciencedirect.com/science/article/pii/S0140673617314630

5. WHO. Measles and Rubella Global Strategic Plan 2012-2020 Midterm Review [Internet]. 2015. Available from: https://www.who.int/immunization/sage/meetings/2016/october/1_MTR_Report_Final_Color_Sept_20_v2.pdf

6. WHO. Measles, fact sheet [Internet]. 2019. Available from: https://www.who.int/news-room/fact-sheets/detail/measles

7. WHO. Joint News Release:More than 140,000 die from measles as cases surge worldwide [Internet]. 2019. Available from: https://www.who.int/news/item/05-12-2019-more-than-140-000-die-from-measles-as-cases-surge-worldwide

8. WHO. Global measles and rubella strategic plan: 2012. 2012;

9. Masresha B, Luce R, Shibeshi M, Katsande R, Fall A, Okeibunor J, et al. Status of measles elimination in eleven countries with high routine immunisation coverage in the WHO African region. J Immunol Sci. 2018;140.

10. Hong H, Makhathini L, Mashele M, Malfeld S, Motsamai T, Sikhosana L, et al. Annual measles and rubella surveillance review, South Africa, 2017. Natl Inst Commun Dis Public Heal Surveill Bull. 2017;16(2):64–77.

11. McMorrow ML, Gebremedhin G, Van den Heever J, Kezaala R, Harris BN, Nandy R. Measles outbreak in South Africa, 2003-2005. South African Med J. 2009;99(5).

12. Ntshoe GM, McAnerney JM, Archer BN, Smit SB, Harris BN, Tempia S, et al. Measles outbreak in South Africa: epidemiology of laboratory-confirmed measles cases and assessment of intervention, 2009-2011. PLoS One [Internet]. 2013/02/20. 2013;8(2):e55682–e55682. Available from: https://pubmed.ncbi.nlm.nih.gov/23437059

13. Hong HA E. Annual measles, rubella and congenital rubella survellance review, South Africa, 2019. 2020; Available from: https://www.nicd.ac.za/wp-content/uploads/2020/06/ANNUAL-MEASLES-RUBELLA-AND-CONGENITAL-RUBELLA-SURVEILLANCE-REVIEW-SOUTH-AFRICA-2019.pdf 18(1).

14. WHO. Experts caution against stagnation of immunization coverage in Africa [Internet]. 2019. Available from: https://www.afro.who.int/news/experts-caution-against-stagnation-immunization-coverage-africa

15. del Mar Mosquera M, Echevarría JE, Puente S, Lahulla F, de Ory F. Use of whole blood dried on filter paper for detection and genotyping of measles virus. J Virol Methods. 2004;117(1):97–9.

16. Hübschen JM, Bork SM, Brown KE, Mankertz A, Santibanez S, Mamou M Ben, et al. Challenges of measles and rubella laboratory diagnostic in the era of elimination. Clin Microbiol Infect. 2017;23(8):511–5.

17. statistics south africa. Mid-Year Population Estimates 2020 [Internet]. Pretoria; 2020. Available from: http://www.statssa.gov.za/?p=13453

18. Massyn N, Day C, Ndlovu N, Padayachee T. District Health Barometer 2019/20. 2020; Available from: https://www.hst.org.za/publications/DistrictHealthBarometers/DHB2019-20CompleteBook.pdf

19. UNICEF. Uneven routine immunization coverage threatens the health of South Africa’s youngest children [Internet]. 2021. Available from: https://www.unicef.org/southafrica/press-releases/uneven-routine-immunization-coverage-threatens-health-south-africas-youngest

20. Hiebert J, Zubach V, Charlton CL, Fenton J, Tipples GA, Fonseca K, et al. Evaluation of Diagnostic Accuracy of Eight Commercial Assays for the Detection of Measles Virus-Specific IgM Antibodies. J Clin Microbiol. 2021;59(6):e03161–20.

21. Thomas HIJ, Barrett E, Hesketh LM, Wynne A, Morgan-Capner P. Simultaneous IgM reactivity by EIA against more than one virus in measles, parvovirus B19 and rubella infection. J Clin Virol. 1999;14(2):107–18.

22. Demicheli V, Rivetti A, Debalini MG, Di Pietrantonj C. Vaccines for measles, mumps and rubella in children. Evidence-Based Child Heal A Cochrane Rev J. 2013;8(6):2076–238.

23. Di Pietrantonj C, Rivetti A, Marchione P, Debalini MG, Demicheli V. Vaccines for measles, mumps, rubella, and varicella in children. Cochrane Database Syst Rev. 2020;(4).

24. Lochlainn LMN, de Gier B, van der Maas N, van Binnendijk R, Strebel PM, Goodman T, et al. Effect of measles vaccination in infants younger than 9 months on the immune response to subsequent measles vaccine doses: a systematic review and meta-analysis. Lancet Infect Dis. 2019;19(11):1246–54.

